# Impact of COVID-19 vaccination on long COVID: a systematic review and meta-analysis

**DOI:** 10.1101/2022.06.20.22276621

**Authors:** Oyungerel Byambasuren, Paulina Stehlik, Justin Clark, Kylie Alcorn, Paul Glasziou

## Abstract

**Background:** The impact of COVID-19 vaccination on preventing or treating long COVID is unclear. We aim to assess the impact of COVID vaccinations administered (i) before and (ii) after acute COVID-19, including vaccination after long COVID diagnosis, on the rates or symptoms of long COVID.

**Methods:** We searched PubMed, Embase, Cochrane COVID-19 trials, and Europe PMC for preprints from 1 Jan 2020 to 16 Feb 2022. We included trials, cohort, and case control studies reporting on long COVID cases and symptoms with vaccine administration both before and after COVID-19 diagnosis as well as after long COVID diagnosis. Risk of bias was assessed using ROBINS-I.

**Results:** We screened 356 articles and found no trials, but 6 observational studies from 3 countries (USA, UK, France) that reported on 442,601 patients. The most common long COVID symptoms studied include fatigue, cough, loss of smell, shortness of breath, loss of taste, headache, muscle ache, trouble sleeping, difficulty concentrating, worry or anxiety, and memory loss or confusion. Four studies reported data on vaccination before SARS-CoV-2 infection, of which three showed statistically significant reduction in long COVID: the odds ratio of developing long COVID with one dose of vaccine ranged between OR 0.22 to 1.03; with two doses OR 0.51 to 1; and with any dose OR 0.85 to 1.01. Three studies reported on post-infection vaccination with odds ratios between 0.38 to 0.91. The high heterogeneity between studies precluded any meaningful meta-analysis. Studies failed to adjust for potential confounders such as other protective behaviours, and missing data, thus increasing the risk of bias, and decreasing the certainty of evidence to low.

**Discussion:** Current studies suggest that COVID-19 vaccinations may have protective and therapeutic effects on long COVID. However, more robust comparative observational studies and trials are urgently needed to clearly determine effectiveness of vaccines in prevention and treatment of long COVID.

## Introduction

Long COVID, also known as post-acute COVID sequalae or post-acute COVID-19 syndrome, is now recognized as a major concern following COVID-19 infections, and likely to cause significant global morbidity for many years to come (1, 2). With global case numbers over 500 million and with a conservative prevalence of 20-30%, more than 100 million people could be affected by long COVID worldwide (3-5).

COVID-19 vaccination can work on three levels to prevent or treat long COVID (Figure 1). Firstly, by preventing infection with SARS-CoV-2 (Level 1); secondly, by reducing the severity of the disease in vaccinated but infected individuals (Level 2); and finally, vaccination may benefit people who already have long COVID (Level 3). However, recent observational studies provide contradictory results and contain methodological flaws which preclude firm conclusions on level 2 and 3 vaccine impact (6, 7).

**Figure 1:**
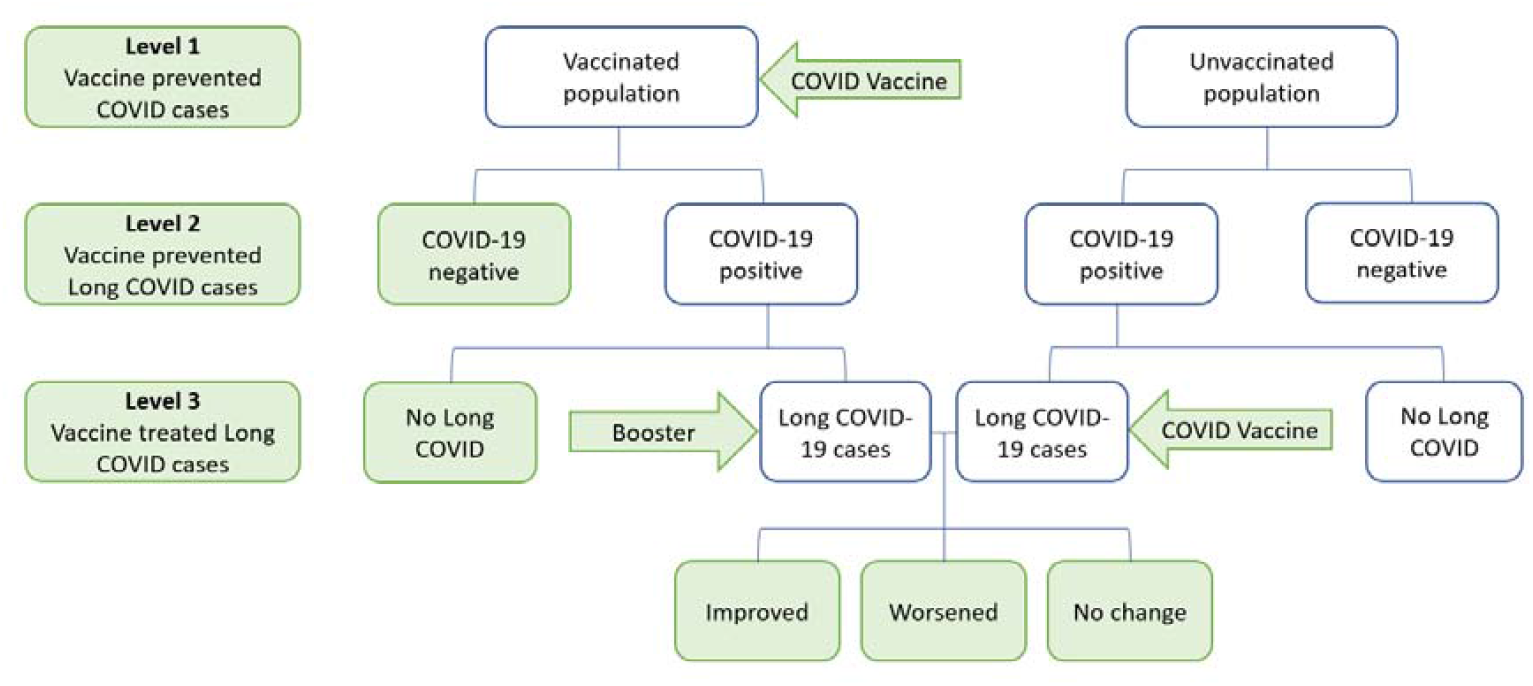
Scope of research question diagram.

Therefore, we aim to assess the impact of COVID vaccinations administered both before and after acute COVID-19, including vaccination after long COVID diagnosis, on the rates or symptoms of long COVID.

## Methods

We conducted a systematic review and a meta-analysis using enhanced processes and automation tools (8). We searched PROSPERO and Open Science Framework (OSF) databases to rule out existence of similar reviews then searched PubMed, Embase, Cochrane COVID-19 trials for published studies, and Europe PMC for pre-prints from 1 January 2020 to 16 Feb 2022. A search string composed of MeSH terms and words was developed in PubMed and was translated to be run in other databases using the Polyglot Search Translator (9). Search strategies for all databases is provided in Supplement 1.

We also conducted forward and backward citation searches of the included studies. For registered studies, we searched ClinicalTrials.gov and World Health Organization – International Clinical Trials Registry Platform (ICTRP). Searches were run from inception to 16 Feb 2022 (see Appendix 1). No publication type or language restrictions were applied. We also contacted authors of large vaccine trials if they had any unpublished data on long-COVID. Our protocol was shared on Open Science Framework (https://osf.io/e8jdy). Ethics approval was not required for this study.

We included randomised controlled trials, cohort studies (retrospectively or prospectively assembled), interrupted time series, and or case-control studies. We excluded case reports, case-series, cross-sectional, and modelling studies. We searched for studies that assessed vaccination status and the emergence of long COVID (history of confirmed or probable COVID-19 within the last 3 months and symptoms that last at least 2 months that cannot be explained by an alternative diagnosis). Studies conducted in community, primary care, and hospital settings were included.

Our inclusion criteria were

- Participants: People of all ages who were eligible to receive COVID vaccination
- Interventions: Any dose of any approved COVID vaccines as recognized by the World Health Organization (WHO) (i.e., BNT162b2 (tozinameran; Pfizer– BioNTech), mRNA-1273 (elasomeran; Moderna), ChAdOx1 nCoV-19 (Oxford– AstraZeneca)), and Ad26.COV2.S (Janssen or Johnson & Johnson) either before the first SARS-CoV-2 infection or after, and after acquisition of long COVID (Figure 1).
- Comparators: No vaccination, active non-COVID-vaccine control (e.g. influenza vaccine), or placebo
- Outcomes: Primary outcome was cases diagnosed with long COVID preferably by health professionals and as defined by the WHO: history of confirmed or probable COVID-19 within the last 3 months and symptoms that last at least 2 months that can’t be explained by an alternative diagnosis (10); Secondary outcomes were prevalence of individual symptoms of long COVID, such as prolonged fatigue, shortness of breath, cognitive difficulties, and anosmia.

We excluded protocols, studies that did not report long COVID outcomes, and studies with uncertain vaccination status (Figure 2).

**Figure 2.**
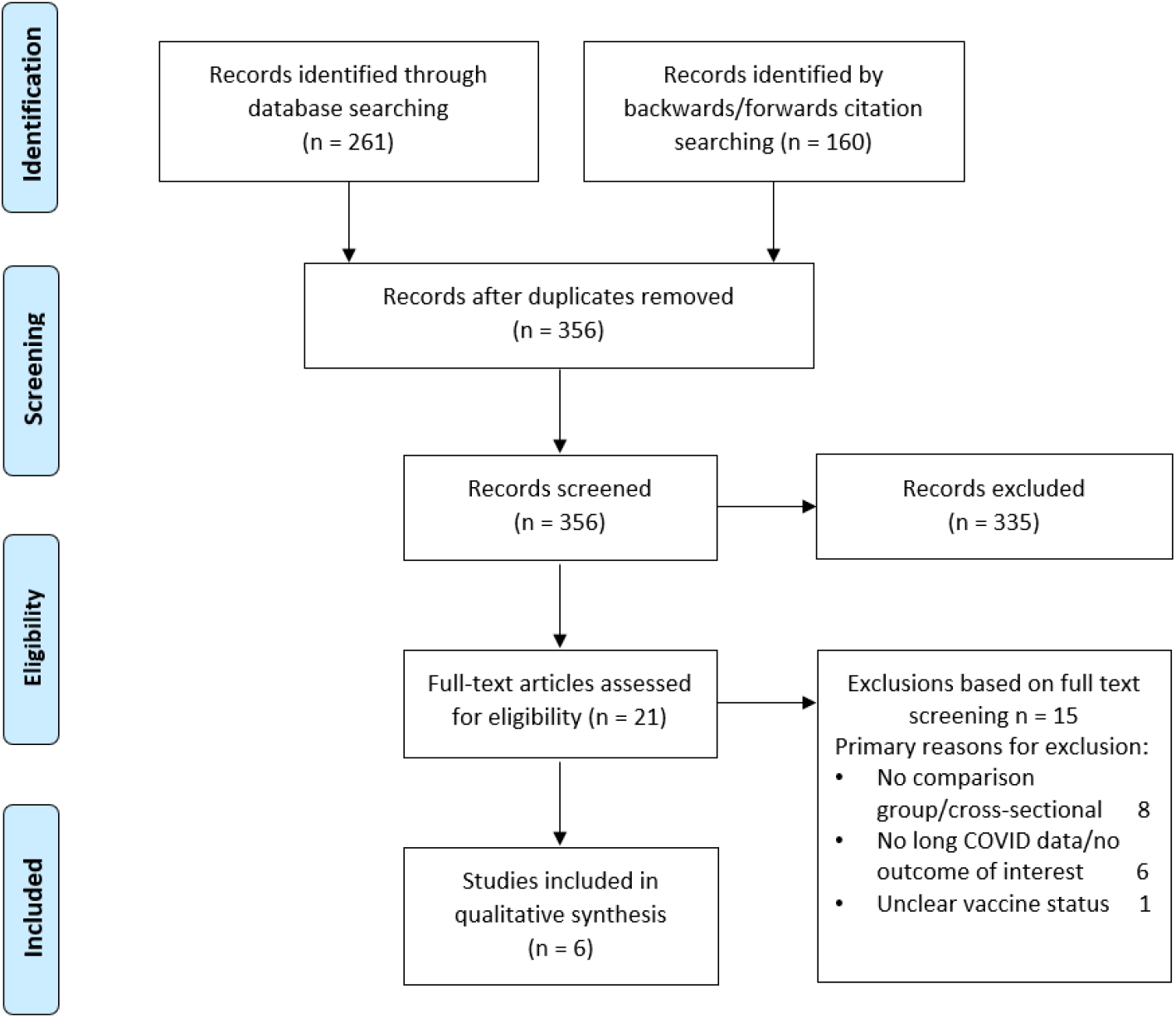
Screening and selection of articles.

### Study selection and screening

Two authors independently (OB, PS) screened titles and abstracts, and full text articles were retrieved for the potentially eligible articles. Two authors (OB, PS) reviewed the full texts against the inclusion criteria. Discrepancies were resolved by referring to a third author (PG). The screening process is summarised in Figure 2. Excluded articles and reasons for exclusion are listed in Supplement 2.

### Data extraction

Data extraction was conducted by two authors (OB, PS) using Microsoft Excel. The following data for study characteristics and outcomes were extracted from each included study:

1. Methods: study authors, year, country, study design, duration of follow-up, setting
2. Participants: number of participants, age, gender, any co-comorbidity
3. Interventions (type of intervention, dose, frequency) and type of comparators (no treatment, other non-COVID vaccine, placebo)
4. Outcomes: cases of long-COVID (primary outcome), and prevalence of individual symptoms (secondary outcome).

### Assessment of risk of bias

Risk of bias was assessed using the ROBINS-I tool, which can assess both randomized and non-randomised studies on a common template (11). Two authors independently assessed risk of bias for each study (OB, PS).

### Data analysis

For dichotomous outcomes, intervention effect was calculated using odds ratios. We did not undertake meta-analyses due to high heterogeneity. We used individual participants as the unit of analysis. Where data were missing or unclear, study investigators were contacted. There were no registered trials for vaccines and long COVID. We were only able to present subgroups by vaccine dose and timing.

## Results

Of 356 titles and abstracts screened, 21 full text articles were assessed for inclusion (Figure 2). There were no eligible randomized trials. The 6 eligible observational studies – 3 preprints and 3 published – were based on data from 3 countries (USA 3, UK 2, France 1) that included 442,601 patients (12-17) (Table 1). Excluded articles and reasons for exclusion are listed in Supplement 2.

**Table 1.** Characteristics of included studies.

| Study ID | Study type and timeframe | Patient or data source | Intervention population | Intervention | Comparator | Outcomes and follow up duration |
| --- | --- | --- | --- | --- | --- | --- |
| **COVID vaccination BEFORE infection** |  |  |  |  |  |  |
| **Al-Aly 2022**<br>(12)<br>(USA)<br>published | Retrospectively assembled cohort.<br>1 Jan - 1 Dec 2021 | US Veterans Health Administration electronic health databases | n=33,940 patients fully vaccinated $\geq 14$ days before positive COVID-19 test, who were still alive 30 days after test.<br>Mean age 67; 9% female | 1 dose of Johnson&Johnson, or 2 doses of Moderna or Pfizer | n=113,474 patients alive 30 days after positive COVID-19 test and no vaccination<br>Mean age 68; 8% female | Risk of at least one post-acute sequelae at 6 months |
| **Antonelli 2022**<br>(13)<br>(UK)<br>published | Prospective, community-based, nested, case-control study. 8 Dec 2020 - 4 Jul 2021 | COVID Symptom Study app users | n=3,071 adult app users with +COVID-19 test $\geq 14$ days after their first vaccine or at least 7 days after their second vaccine; and had no positive test before vax | 1<sup>st</sup> or 2<sup>nd</sup> dose of Pfizer, Moderna, and AstraZeneca | n=3,244 unvaccinated participants reporting a positive SARS-CoV-2 test who had used the app for at least 14 days after the test | Long-duration ($\geq 1$ month) symptoms following 1 dose. Most common symptoms. |
| **Simon 2021**<br>(16)<br>(USA)<br>preprint | Retrospectively assembled cohort.<br>Feb 2020 - May 2021 | Arcadia Data Research dataset with > 150 million patients records | n=2,392 patients got first dose before COVID-19 diagnosis; 61% female | 1 dose of Pfizer-BioNTech, Moderna, or Johnson & Johnson | n=220,460 patients with no vaccination before COVID and 12 weeks after.<br>60% female | Odds of having long-COVID. 3 months |
| **Taquet 2021**<br>(14)<br>(USA)<br>preprint | Retrospectively assembled cohort.<br>1 Jan – 31 Aug 2021 | TriNetX Analytics – federated network of linked EHR with 81 million patient data | n=9,479 adults who received a COVID-19 vaccine at least 2 weeks before SARS-CoV-2 infection, mean age 57, 60% female | Pfizer, Moderna, or Johnson&Johnson | n=9,479 propensity matched pts with Flu vax at any time, mean age 58, 61% female | For any long COVID feature within 6 months |
| **COVID vaccination AFTER infection or after Long COVID diagnosis** |  |  |  |  |  |  |
| **Ayoubkhani 2022**<br>(15)<br>(UK)<br>published | Interrupted time-series.<br>3 Feb - 5 Sep 2021 | COVID-19 Infection Survey participants | n=28,356 patients with long COVID, at least 1 vaccination after diagnosis.<br>mean age 46 years; 56% female, | AstraZeneca, Pfizer, Moderna | Self-controlled (symptoms before vaccine) | Long COVID of any severity following 1st and 2nd dose $\geq 3$ months. 10 most common symptoms. |
| **Simon 2021**<br>(16)<br>(USA)<br>preprint | Retrospectively assembled cohort.<br>Feb 2020 - May 2021 | Arcadia Data Research dataset with > 150 million patients records | n=17,796 patients diagnosed with COVID-19, either by PCR or by ICD-10 code U07.1 at any time or B97.29 prior to May 2020 and vaccinated within 12 weeks after COVID-19 diagnosis; 61% female | 1 dose of Pfizer-BioNTech, Moderna, or Johnson & Johnson | Same as above | Odds of having long-COVID. 3 months |
| **Tran 2021**<br>(17)<br>(France)<br>preprint | Prospective cohort (target trial emulation) Dec 2020 - Sep 2021 | ComPaRe (cohort of chronic disease sufferers) | n=455 adults with a confirmed or suspected COVID-19 infection and at least one symptom attributable to long COVID reported at baseline and persisting more than 3 weeks. Median age 47; 81% female | AstraZeneca, Pfizer, Moderna, or Johnson&Johnson | n=455 unvaccinated control 1:1 propensity matched people from the same cohort | Remission of all long COVID symptoms by 4 months |

Three studies assessed effect of vaccination given before SARS-CoV-2 infection (Level 2) (12-14); one study assessed vaccine effects after infection, and two studies were of vaccination given to people with long COVID (Level 3) (15-17). One study provided data for both vaccination before and after infection and therefore was entered twice in Table 1 (16). Four of the studies used data from large medical databases (12, 14-16), one study used COVID symptom study app user data (13), and one study recruited patients who already had long COVID symptoms to prospectively follow (17).

All the studies were conducted and concluded by Nov 2021 and therefore did not use the current long COVID case definition from WHO. In three studies, the long COVID diagnosis were self-made or reported by patients (13, 15, 17) and the others used electronic health record data to extract long COVID symptoms as a proxy for the diagnosis (12, 14, 16).

Secondary outcome was reported in two studies (13, 15). The most common long COVID symptoms include fatigue, cough, weakness and tiredness, loss of smell, shortness of breath, loss of taste, headache, trouble sleeping, difficulty concentrating, muscle ache, worry or anxiety, and memory loss or confusion.

### Effect of vaccination on long COVID outcomes

The high heterogeneity between studies precluded a meaningful meta-analysis. The forest plot of each study outcomes shows high statistical heterogeneity (Figure 3). Four studies reported data on vaccination before SARS-CoV-2 infection (12-14, 16), of which three showed statistically significant reduction in long COVID (12, 13, 16). The odds ratio (OR) of developing long COVID with one dose of pre-infection vaccination ranged from 0.22 to 1.03; for two doses the OR ranged from 0.51 to 1; and with any dose before infection the OR ranged from 0.85 to 1.01. The 3 studies that reported data on post-infection vaccination had ORs ranging from 0.38 to 0.91 (all statistically significant) (15-17).

**Figure 3.**
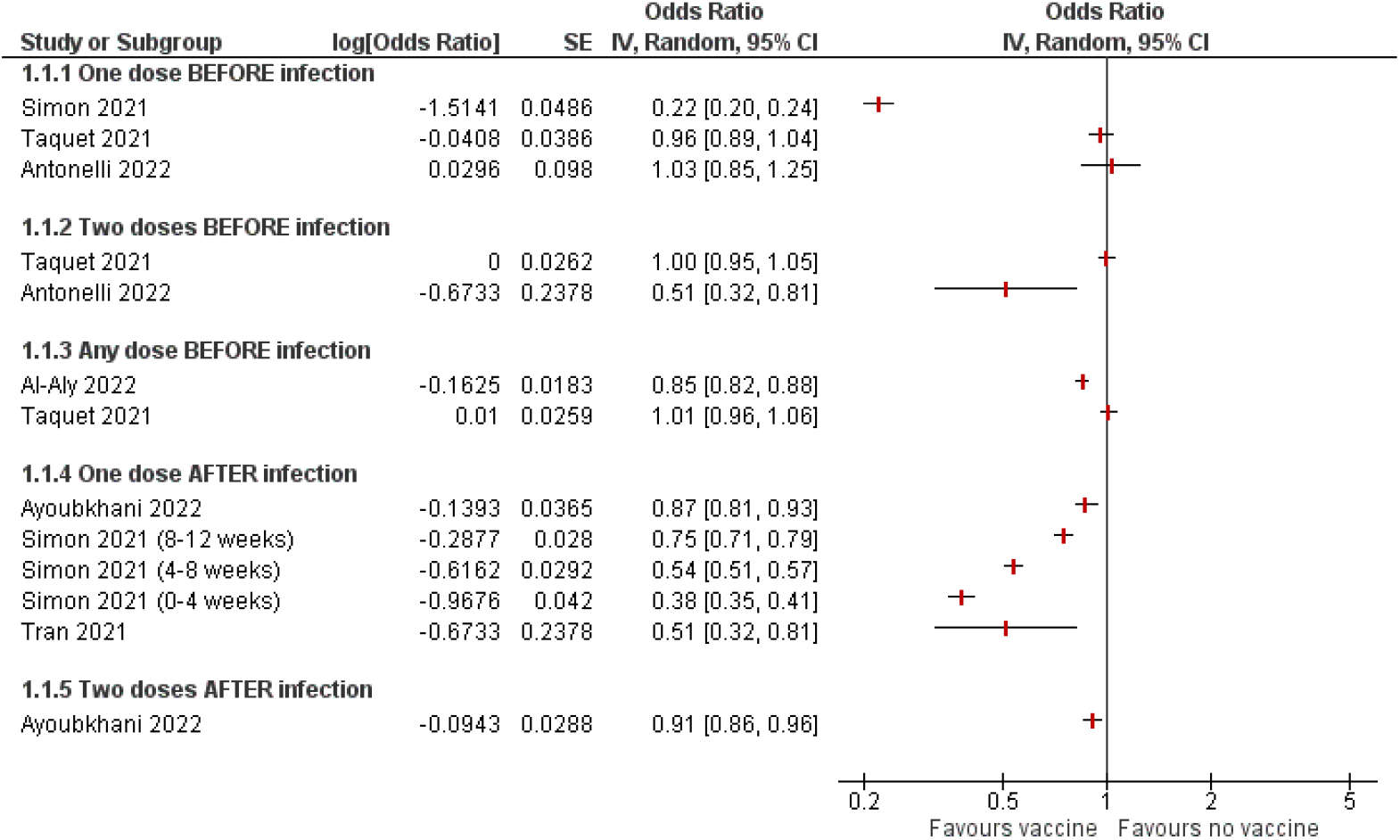
Impact of vaccinations on long COVID forest plot.

### Risk of bias in included studies

The risk of bias of the included studies was assessed by ROBINS-I for non-randomised studies of interventions. The risk of bias of the individual studies was judged overall as low to moderate, with confounding and bias due to missing data being the primary sources of bias. Main issues arising from confounding were not accounting for vaccine hesitancy or original disease severity. Most of the studies did not report on data missingness or how missing data was dealt with. Risk of selection bias, classification of intervention, measurement of outcomes and selection of reported results were judged to be low risk of bias (Table 2).

**Table 2:** Risk of bias in included studies assessed by ROBINS-I.

| Risk of bias domains<br>Study ID | Bias due to confounding | Bias in selection of participants | Bias in classification of interventions | Bias due to deviations from intended interventions | Bias due to missing data | Bias in measurement of outcomes | Bias in selection of the reported result | Overall risk of bias |
| --- | --- | --- | --- | --- | --- | --- | --- | --- |
| COVID vaccination before infection |  |  |  |  |  |  |  |  |
| Al-Aly 2022 | Low | Low | Low | Low | Low | Low | Low | Low |
| Antonelli 2022 | Moderate | Low | Low | Low | Moderate | Low | Low | Moderate |
| Taquet 2021 | Low | Low | Low | Low | Moderate | Low | Low | Moderate |
| COVID vaccination after infection or after Long COVID diagnosis |  |  |  |  |  |  |  |  |
| Ayoubkhani 2022 | Low | Low | Low | Low | Moderate | Low | Low | Moderate |
| Simon 2021 | Serious | Low | Low | Low | Moderate | Low | Low | Moderate |
| Tran 2021 | Low | Low | Low | Low | Low | Moderate | Low | Low |

## Discussion

### Principal findings

Of the six observational studies identified, all but one showed statistically significant reductions in long COVID rates with both pre- and post-infection vaccination. There was not enough data to examine any dose-response relationship. All six studies were non-randomized, and most did not adjust for missing data, so the overall risk of bias was moderate. Thus, the evidence summarised here is of low certainty.

### Strengths and weaknesses of the study

Strengths of our review include the search of multiple databases for published, preprint, and unpublished articles including public health reports. We critically assessed risk of bias of most full text articles we screened to ascertain to include studies with the least risk of bias.

However, there are several limitations to our findings. Although we set out to explore long COVID symptomatology as a secondary outcome, not many of our included studies reported it. There were several studies that showed symptom changes following vaccination, however they were mostly cross-sectional in nature and thus impossible to establish true causality, and thus excluded. Furthermore, long COVID characteristics and symptomatology are getting well established with global data (1, 5, 18).

### Strengths and weaknesses in relation to other studies

Two government reports - by Public Health Ontario and UK Health Security agency - aimed to estimate vaccine impact on long COVID (19, 20). Both were done as rapid reviews; thus, did not conduct a rigorous search nor quality assessments on the reported studies. They included multiple cross-sectional studies and only narratively explained the findings. Due to the lack of rigorous inclusion criteria, these reviews cannot be used to establish effectiveness of vaccines to prevent long COVID.

### Meaning of the study

Vaccine for COVID-19 has been found to prevent cases, particularly for the earlier variants in the pandemic and hence would prevent long COVID by preventing initial infection (Level 1, Figure 1). Less clear, though highly plausible, has been whether vaccines – by reducing the severity of COVID-19 (Level 2, Figure 1) reduce long COVID following infection. The studies identified here are inconsistent, though the study with lowest risk of bias shows an important and statistically significant reduction. Vaccination after infection and in those with long COVID has been more controversial, but the studies here are reassuringly consistent in being protective.

### Unanswered questions and future research

A key finding from this review is the paucity of good studies, in particular randomized trials, to address the impact of vaccines on long COVID. This has several implications for future research. First, the ideal data on vaccine’s impact on long COVID cases following breakthrough cases could have come from large clinical trials of vaccines. However, our search into such data revealed vaccine efficacy trials did not plan and or collect suitable data for such outcome. However, it may still be possible to design follow up studies of breakthrough cases from the large vaccine trials to estimate rates of long COVID.

Second, ongoing vaccine effectiveness trials on children should include provisions for longer follow up on the breakthrough cases and post-infection vaccination cases. Third, all our included studies were conducted before Nov 2021, thus do not include data from Omicron variant wave. UK Office for National Statistics data shows Omicron variant has caused the greatest number of COVID and long COVID cases in the UK (21). But a new analysis that compared delta and omicron periods in the UK shows long COVID prevalence is twice as less in omicron variant waves and omicron cases were less likely to experience long COVID even with more than 6 months gap between vaccination and infection (OR 0.24 to 0.50 (22). Mapping long COVID data with all the different subvariants of SARS-CoV-2 will also help inform the public health measures against the pandemic spread. Short of that, researchers should employ trial emulation techniques to better estimate the effect of vaccines on different age groups and variants. In our review, only one study explicitly emulated a target trial (14) and two others used propensity score matching when creating their comparator cohorts (12, 17).

Fourth, the data from our included studies also suggest that COVID vaccinations at least provide equipoise in terms of long COVID prevention and or treatment, thus, trials on vaccination effect for post-infectious and post-long COVID patients should be conducted as a priority. Finally, medical professionals’ awareness of long COVID case definition and management in parallel with long COVID patients’ care needs should be explored.

COVID-19 vaccinations have saved millions of lives and severe cases. However, the impact of vaccines on preventing or treating long COVID could not have been conclusively established in this review. Many unanswered questions remain and needs to be answered as high priority.

## Key messages

- There are no trials assessing the impact of COVID-19 vaccination on preventing long COVID.
- Data from six observational studies suggest COVID-19 vaccination may protect against long COVID.
- More robust comparative observational studies and trials are urgently needed to clearly determine effectiveness of vaccines in prevention and treatment of long COVID.
- Any new vaccine trial should have adequate follow up to assess long COVID as an outcome.

## Data Availability

All data produced in the present work are contained in the manuscript

## Authors’ contributions

PG conceived the study and co-designed with OB, PS, and KA. JC led the literature searches including backward and forward citation analysis. OB and PS conducted the parallel title, abstract and full text screening. OB and PS did data extraction and analysis. All authors contributed to resolving disagreements throughout the study conduct and to writing of the manuscript. PG is guarantor for the article.

## Acknowledgments

We thank the authors of eligible manuscripts for their replies to our queries. We also thank Professor David Henry for his methodological expertise on the risk of bias assessment.

