## Supplementary material for "Impact of COVID-19 vaccination on long COVID: a systematic review and meta-analysis"

### Supplementary material for the article “Impact of COVID-19 vaccination on long COVID: a systematic review and meta-analysis”

#### Supplement 1. Search Strategies

##### PubMed - run [no date specified]

**PubMed**

("Post-acute COVID-19 syndrome"[NM] OR "COVID-19/complications"[Mesh] OR "Long covid"[tiab] OR Long-covid[tiab] OR "COVID-19 sequelae"[tiab] OR "COVID 19 sequelae"[tiab] OR "Post-acute COVID-19"[tiab] OR "Post Covid"[tiab] OR ((Post-vaccination[tiab] OR Post vaccination[tiab]) AND (SARS-CoV-2[tiab] OR Covid-19[tiab] OR "Covid 19"[tiab])))

AND

("COVID-19 Vaccines"[Mesh] OR "Vaccination"[Mesh] OR Vaccination[tiab] OR Vaccinated[tiab] OR Vaccines[tiab] OR Inoculated[tiab] OR Inoculation[tiab])

AND

(Symptoms[tiab] OR Symptom[tiab] OR "Post-acute outcomes"[tiab] OR "Post acute outcomes"[tiab] OR Long-term[tiab] OR "Long term"[tiab])

AND

("Morbidity"[Mesh] OR Epidemiology[sh] OR Incidence[tiab] OR Incidences[tiab] OR Prevalence[tiab] OR Trajectory[tiab] OR Persistent[tiab] OR "Risk factor"[tiab])

##### The Cochrane Library for clinical trials in CENTRAL - run [no date specified]

**Cochrane CENTRAL**

("Post-acute COVID-19 syndrome":kw OR "Long covid":ti,ab OR "COVID 19 sequelae":ti,ab OR "COVID 19 sequelae":ti,ab OR "Post acute COVID 19":ti,ab OR "Post Covid":ti,ab OR ((Post vaccination:ti,ab OR "Post vaccination":ti,ab) AND (SARS CoV 2:ti,ab OR "Covid 19":ti,ab)))

AND

([mh "COVID-19 Vaccines"] OR [mh Vaccination] OR Vaccination:ti,ab OR Vaccinated:ti,ab OR Vaccines:ti,ab OR Inoculated:ti,ab OR Inoculation:ti,ab)

AND

(Symptoms:ti,ab OR Symptom:ti,ab OR "Post-acute outcomes":ti,ab OR "Post acute outcomes":ti,ab OR Long-term:ti,ab OR "Long term":ti,ab)

AND

([mh Morbidity] OR [mh /EP] OR Incidence:ti,ab OR Incidences:ti,ab OR Prevalence:ti,ab OR Trajectory:ti,ab OR Persistent:ti,ab OR "Risk factor":ti,ab)

#### Embase via Elsevier - run [no date specified]

##### Embase

('long COVID'/exp OR "Long covid":ti,ab OR Long-covid:ti,ab OR "COVID-19 sequelae":ti,ab OR "COVID 19 sequelae":ti,ab OR "Post-acute COVID-19":ti,ab OR "Post Covid":ti,ab OR ((Post-vaccination:ti,ab OR "Post vaccination":ti,ab) AND (SARS-CoV-2:ti,ab OR Covid-19:ti,ab OR "Covid 19":ti,ab)))

AND

("SARS-CoV-2 vaccine"/exp OR Vaccination/exp OR Vaccination:ti,ab OR Vaccinated:ti,ab OR Vaccines:ti,ab OR Inoculated:ti,ab OR Inoculation:ti,ab)

AND

(Symptoms:ti,ab OR Symptom:ti,ab OR "Post-acute outcomes":ti,ab OR "Post acute outcomes":ti,ab OR Long-term:ti,ab OR "Long term":ti,ab)

AND

(Morbidity/exp OR "Epidemiology":ti,ab OR Incidence:ti,ab OR Incidences:ti,ab OR Prevalence:ti,ab OR Trajectory:ti,ab OR Persistent:ti,ab OR "Risk factor":ti,ab)

#### EuropePMC (preprints) - run [no date specified]

##### Preprints – via Europe PMC

(TITLE:"Long covid" OR TITLE:Long-covid OR ABSTRACT:"Long covid" OR ABSTRACT:Long-covid OR TITLE:"COVID-19 sequelae" OR ABSTRACT:"COVID-19 sequelae")

AND

(TITLE:Vaccination OR TITLE:Vaccination OR TITLE:Vaccinated OR TITLE:Vaccines OR TITLE:Inoculated OR TITLE:Inoculation OR ABSTRACT:Vaccination OR ABSTRACT:Vaccination OR ABSTRACT:Vaccinated OR ABSTRACT:Vaccines OR ABSTRACT:Inoculated OR ABSTRACT:Inoculation)

AND

(TITLE:Symptoms OR ABSTRACT:Symptoms OR TITLE:"Post-acute outcomes" OR ABSTRACT:"Post-acute outcomes" OR TITLE:Incidence OR ABSTRACT:Incidence OR TITLE:Incidences OR ABSTRACT:Incidences OR TITLE:Trajectory OR ABSTRACT:Trajectory OR TITLE:Persistent OR ABSTRACT:Persistent)

#### Supplement 2. Excluded studies following full text screening with reasons

| No. | Excluded articles | Reason |
|---|---|---|
| 1. | Arjun MC, Singh AK, Pal D, Das K, Gajjala A, Venkateshan M, et al. Prevalence, characteristics, and predictors of Long COVID among diagnosed cases of COVID-19. medRxiv. 2022:2022.01.04.21268536. | Cross sectional |
| 2. | Arnold DT, Milne A, Samms E, Stadon L, Maskell NA, Hamilton FW. Are vaccines safe in patients with Long COVID? A prospective observational study. 2021. | No outcome of interest |
| 3. | Gaber TA-ZK, Ashish A, Unsworth A, Martindale J. Are mRNA Covid 19 vaccines safe in Long Covid patients? A Health Care Workers perspective. British Journal of Medical Practitioners. 2021;14(1). | No comparison group |
| 4. | Herman B, Viwattanakulvanid P, Dzulhadj A, Oo AC, Patricia K, Pongpanich S. Effect of full vaccination and post-covid olfactory dysfunction in recovered COVID-19 patient. A retrospective longitudinal study with propensity matching. medRxiv. 2022:2022.01.10.22269007. | No long COVID outcome of interest |
| 5. | Kuodi P, Gorelik Y, Zayyad H, Wertheim O, Wiegler KB, Jabal KA, et al. Association between vaccination status and reported incidence of post-acute COVID-19 symptoms in Israel: a cross-sectional study of patients tested between March 2020 and November 2021. 2022. | Unclear vaccination status at time of infection |
| 6. | Massey D, Berrent D, Akrami A, Assaf G, Davis H, Harris K, et al. Change in Symptoms and Immune Response in People with Post-Acute Sequelae of SARS-Cov-2 Infection (PASC) After SARS-Cov-2 Vaccination. 2021. | No long COVID data, protocol |
| 7. | Nehme M, Braillard O, Salamun J, Jacquerioz F, Courvoisier DS, Spechbach H, et al. Symptoms After COVID-19 Vaccination in Patients with Post-Acute Sequelae of SARS-CoV-2. Journal of General Internal Medicine. 2022. | Cross sectional |
| 8. | Scherlinger M, Pijnenburg L, Chatelus E, Arnaud L, Gottenberg JE, Sibilia J, et al. Effect of SARS-CoV-2 Vaccination on Symptoms from Post-Acute Sequelae of COVID-19: Results from the Nationwide VAXILONG Study. Vaccines. 2022;10(1). | No comparison group |
| 9. | Senjam S, Singh B, Parmeshwar K, Nichal N, Manna S, Madan K, et al. Assessment of Post COVID-19 Health Problems and its Determinants in North India: A descriptive cross section study. 2021. | Cross sectional |
| 10. | Sheikh A, McMenamin J, Taylor B, Robertson C. SARS-CoV-2 Delta VOC in Scotland: demographics, risk of hospital admission, and vaccine effectiveness. Lancet (London, England). 2021;397:2461-2. | No long COVID data |

| | | |
|---|---|---|
| 11. | Strahm C, Seneghini M, Güsewell S, Egger T, Leal-Neto O, Brucher A, et al. Symptoms Compatible With Long Coronavirus Disease (COVID) in Healthcare Workers With and Without Severe Acute Respiratory Syndrome Coronavirus 2 (SARS-CoV-2) Infection—Results of a Prospective Multicenter Cohort. Clinical Infectious Diseases. 2022. | No outcome of interest |
| 12. | Strain WD, Sherwood O, Banerjee A, van der Togt V, Hishmeh L, Rossman J. The Impact of COVID Vaccination on Symptoms of Long COVID. An International Survey of People with Lived Experience of Long COVID. 2021. | No comparison group |
| 13. | Wanga V, Chevinsky J, Dimitrov L, Gerdes M, Whitfield G, Bonacci R, et al. Long-Term Symptoms Among Adults Tested for SARS-CoV-2 — United States, January 2020–April 2021. 2021. | No comparison group |
| 14. | Whittaker H, Gulea C, Koteci A, Kallis C, Morgan A, Iwundu C, et al. GP consultation rates for sequelae after acute covid-19 in patients managed in the community or hospital in the UK: population based study. BMJ (Clinical research ed). 2021;375:e065834. | No comparison group |
| 15. | Wisnivesky JP, Govindarajulu U, Bagiella E, Goswami R, Kale M, Campbell KN, et al. Association of Vaccination with the Persistence of Post-COVID Symptoms. Journal of General Internal Medicine. 2022. | No outcome of interest |
